# Cryopreserved Peripheral Blood Mononuclear Cells Recover Mitochondrial Function Following Overnight Culture Revealed by High-Resolution Respirometry

**DOI:** 10.64898/2026.11.02.26366883

**Authors:** Robert D. ʻAtiola, Christopher Puliʼuvea, Natalie E. Netzler, Troy L. Merry, Garcia Vailahi, Anthony J. R. Hickey, Peter Shepherd, Christopher P. Hedges

## Abstract

**Background:** High-resolution respirometry of peripheral blood mononuclear cells (PBMCs) is widely used to assess mitochondrial function, but reliance on freshly isolated cells limits flexibility and sample batching. The impact of cryopreservation and post-thaw handling on PBMC mitochondrial respiration remains incompletely defined.

**Methods:** PBMCs from healthy young adults were analysed under three conditions: freshly isolated, cryopreserved and directly thawed, and cryopreserved followed by overnight recovery in supplemented culture medium. Mitochondrial respiration was assessed using an Oroboros O2k high-resolution respirometer following plasma membrane permeabilisation and a substrate–uncoupler–inhibitor titration protocol. Respiration parameters included leak, oxidative phosphorylation (OXPHOS), maximal uncoupled respiration, and non-mitochondrial oxygen consumption.

**Results:** Directly thawed PBMCs exhibited reduced ATP-coupled and maximal respiratory flux compared with fresh cells, indicating acute cryopreservation-induced mitochondrial suppression. In contrast, PBMCs undergoing overnight recovery showed substantial restoration of mitochondrial function, with OXPHOS and uncoupled respiration closely approximating those of fresh cells. Cell viability was comparable across conditions.

**Conclusion:** Cryopreserved PBMCs can be reliably used for mitochondrial respirometry when an overnight recovery step is included. This approach enables sample storage, batching, and broader application in large-scale and retrospective human studies.

## Introduction

Metabolic processes generate the metabolites required for cell survival and proliferation, with mitochondria playing a central role in coordinating these pathways. Oxidative phosphorylation (OXPHOS), which occurs via the electron transport chain (ETC) located in the inner mitochondrial membrane, is essential for ATP production in most mammalian cells [1]. Through this process, energy substrates derived from glucose, amino acids, and lipids are converted into ATP to sustain cellular function. Because oxygen consumption is tightly coupled to ATP production, changes in mitochondrial respiration can directly influence cellular and systemic metabolic function [2].

Mitochondrial dysfunction occurs when mitochondria fail to perform optimally and is often associated with mutations in mitochondrial DNA (mtDNA) or nuclear DNA (nucDNA) encoding mitochondrial components [3-5]. It can also be characterised by increased production of reactive oxygen species, reduced oxidative phosphorylation capacity, and impaired protein assembly [6]. Importantly, growing evidence links mitochondrial dysfunction to a range of metabolic and systemic diseases [7-10].

Accurate assessment of mitochondrial function is therefore critical for understanding both physiological and pathological processes. Over time, various optimised methods have been developed to measure mitochondrial function at both the tissue and cellular levels [11, 12]. However, many of these approaches require freshly isolated samples to ensure reliable measurements. Freeze–thaw processes can disrupt mitochondrial membranes, promote the loss of cytochrome c from the intermembrane space, and uncouple oxygen consumption from ATP synthesis, thereby impairing ETC activity [13]. These limitations present a significant challenge for both basic and translational research, as they restrict the ability to batch samples, reduce costs, and analyse stored specimens.

Recent work by Acín-Pérez et al. (2020) [14] introduced a novel approach to measure mitochondrial respiration in frozen biological samples, demonstrating that respiratory function can be partially preserved under specific experimental conditions. Using murine tissues and human cell lines, the study showed that, when combined with plasma membrane permeabilisation and appropriate substrate–inhibitor conditions, oxygen consumption rates could be maintained across fresh and frozen specimens. However, these findings were obtained using Seahorse XF96 microplate-based assays, and their applicability to high-resolution respirometry in primary human cells remains unclear. Therefore, the aim of this study was to determine whether mitochondrial function measured in cryopreserved human PBMCs reflects that of freshly isolated PBMCs, and to assess whether an overnight recovery period following thawing can restore mitochondrial respiration. Establishing such a workflow would enable more flexible experimental designs, including sample batching, multi-site studies, and retrospective analyses using biobanked specimens

## Methods

### Participant recruitment

Six healthy young participants (n = 3 male, n = 3 female) were recruited for this study. Participants were free from cardiovascular and metabolic disease and from any injuries that would preclude blood donation. Inclusion criteria required no blood donation within the previous 24 h and absence of medications or substances that could influence physiological responses. Written informed consent was obtained prior to participation. The study was approved by the Auckland Human Participants Ethics Committee (reference 010558).

### Sample collection, preparation and cryopreservation

Participants arrived at the laboratory at 0700–0800 after an overnight fast and had abstained from physical activity for at least 48 h before the visit. Approximately 45-50 ml of resting venous blood was collected into K2-EDTA Vacutainer tubes and used to isolate PBMCs. PBMCs were isolated from whole blood by 50 mL SepMate™ tubes (STEMCELL Technologies), which were loaded with 15mL density gradient centrifugation (Ficoll-Paque Plus; GE Healthcare Life Sciences) and centrifuged for 1min to enable the Ficoll to sit below the membrane. 35 mL of diluted blood was then carefully layered onto the membrane, and samples were centrifuged at 1200 x g for 10 minutes, with the brake on at room temperature (RT). Following centrifugation, mononuclear cells were harvested from the sample/medium interface using a Pasteur pipette. Cells were subsequently washed twice in room temperature RPMI-1640-medium (non-supplemented) to remove residual Ficoll. In all cases, isolation of PBMCs commenced within 1 h of the collection of a whole-blood sample. Isolated cells were manually counted and assessed as > 99% viable using trypan blue. Cells for mitochondrial respiration analysis (12–16 million cells total) were pelleted and resuspended in respiration media (MiR05), and PBMC mitochondrial respiration was evaluated 2–3 h after initial blood draw. The remaining PBMCs were snap-frozen and stored at -80°C overnight before being transferred to long-term storage in liquid nitrogen.

PBMC cryopreservation involved first preparing a 2× cryopreservation medium (DMSO + FBS), stored at −20 °C and thawed at 37 °C before use, followed by careful removal of the supernatant to leave the cell pellet intact, then resuspension in culture medium (fully supplemented RPMI-1640-medium (10% FBS, 1% PS, 1% GlutaMAX). The cryopreservation medium, pre-cooled to 4 °C, was then added dropwise at a 1:1 volume ratio to the cell suspension to reach a final 10% DMSO concentration, ensuring gentle mixing and reduced osmotic shock. Cells were aliquoted at 1 mL per cryovial (typically across multiple tubes per donor), with mixing steps such as vortexing to maintain consistency. The cryovials were immediately placed into an isopropanol-filled controlled-rate freezing container (Mr Frosty) and stored at −80 °C overnight before transfer to liquid nitrogen for long-term storage (**Figure 1**). Finally, sample quality was evaluated by post-thaw viability testing compared with fresh cells.

**Figure 1.**
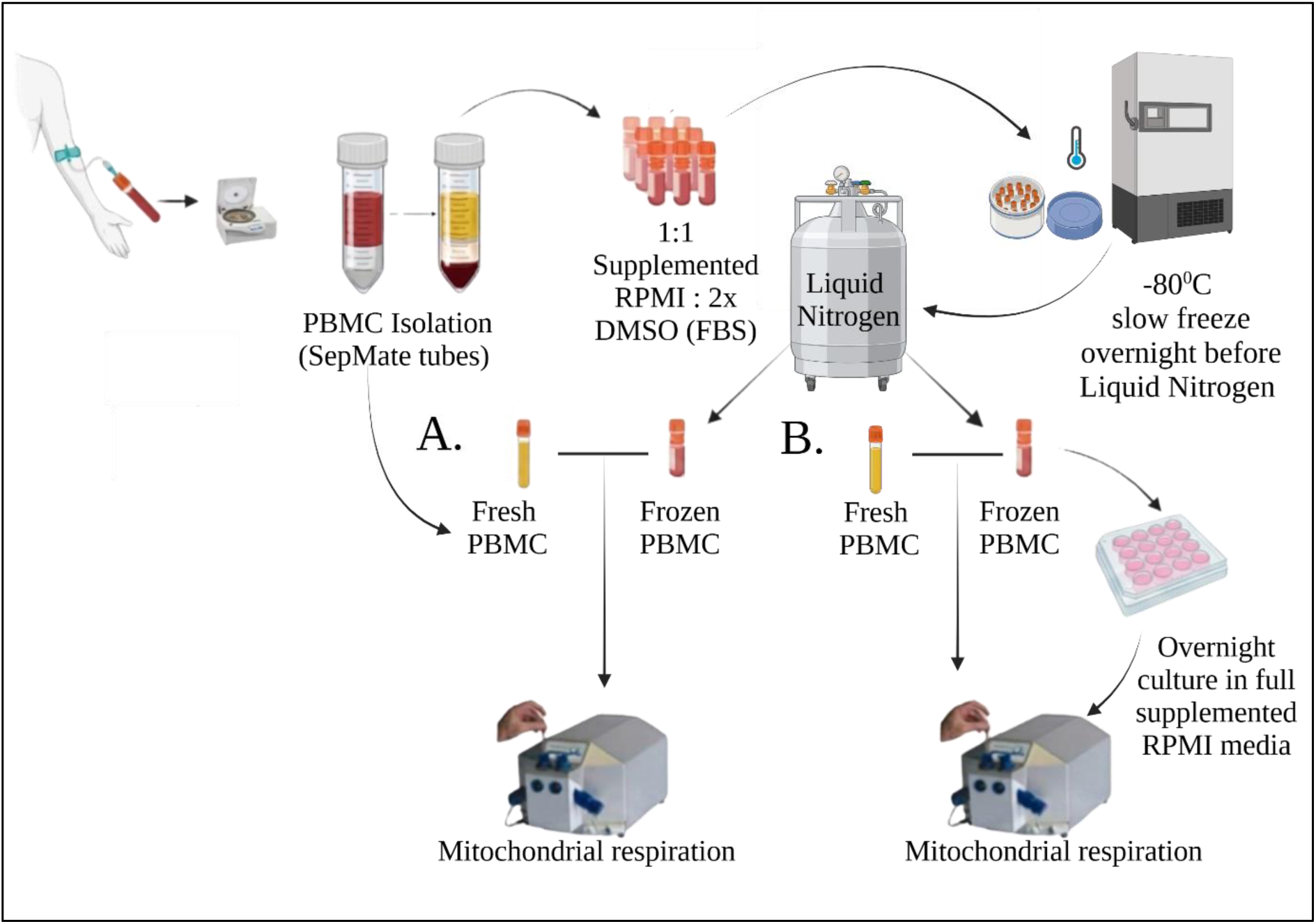
Schematic overview of the experimental design used to assess the effects of cryopreservation and post-thaw recovery on mitochondrial respiration in human peripheral blood mononuclear cells (PBMCs). (A) Comparison between freshly isolated PBMCs assayed immediately after isolation and cryopreserved PBMCs that were rapidly thawed and analysed directly without recovery. (B) Comparison between freshly isolated PBMCs and cryopreserved PBMCs that were thawed and allowed to recover overnight in fully supplemented RPMI prior to analysis. For both panels, mitochondrial respiration was measured using high-resolution respirometry (Oroboros O2k) following plasma membrane permeabilisation with digitonin. A standardized substrate–uncoupler–inhibitor titration protocol was used to quantify leak respiration, oxidative phosphorylation (OXPHOS), maximal uncoupled respiration, and non-mitochondrial oxygen consumption.

### Cell viability and recovery calculations

Cell viability was calculated as:

%viability = (number of viable PBMCs/ number of total PBMCs) x 100

1. Recovery (%) directly after thawing: % Recovery directly after thawing = number of viable PBMCs after thawing/ number of frozen viable PBMCs x 100
2. Recovery (%) after overnight culture: % Recovery after overnight culture = ((number of viable PBMCs after overnight culture)/ (number of frozen viable PBMCs - number of PBMCs removed for measurement directly after thawing)) x 100.

### Cell culture

Cells were maintained at 37 °C in a humidified incubator with 5% CO₂. For overnight recovery experiments, cryopreserved PBMCs were thawed, washed in RPMI 1640 (wash media; Table 1), and resuspended in fully supplemented RPMI (RF10; Table 1: RPMI 1640 with 10% FBS, 1% Penicillin–Streptomycin, and 1% GlutaMAX™). Cells were plated in ultra-low attachment 6-well plates (Corning Costar®) and cultured overnight prior to analysis (Figure 2).

**Figure 2.**
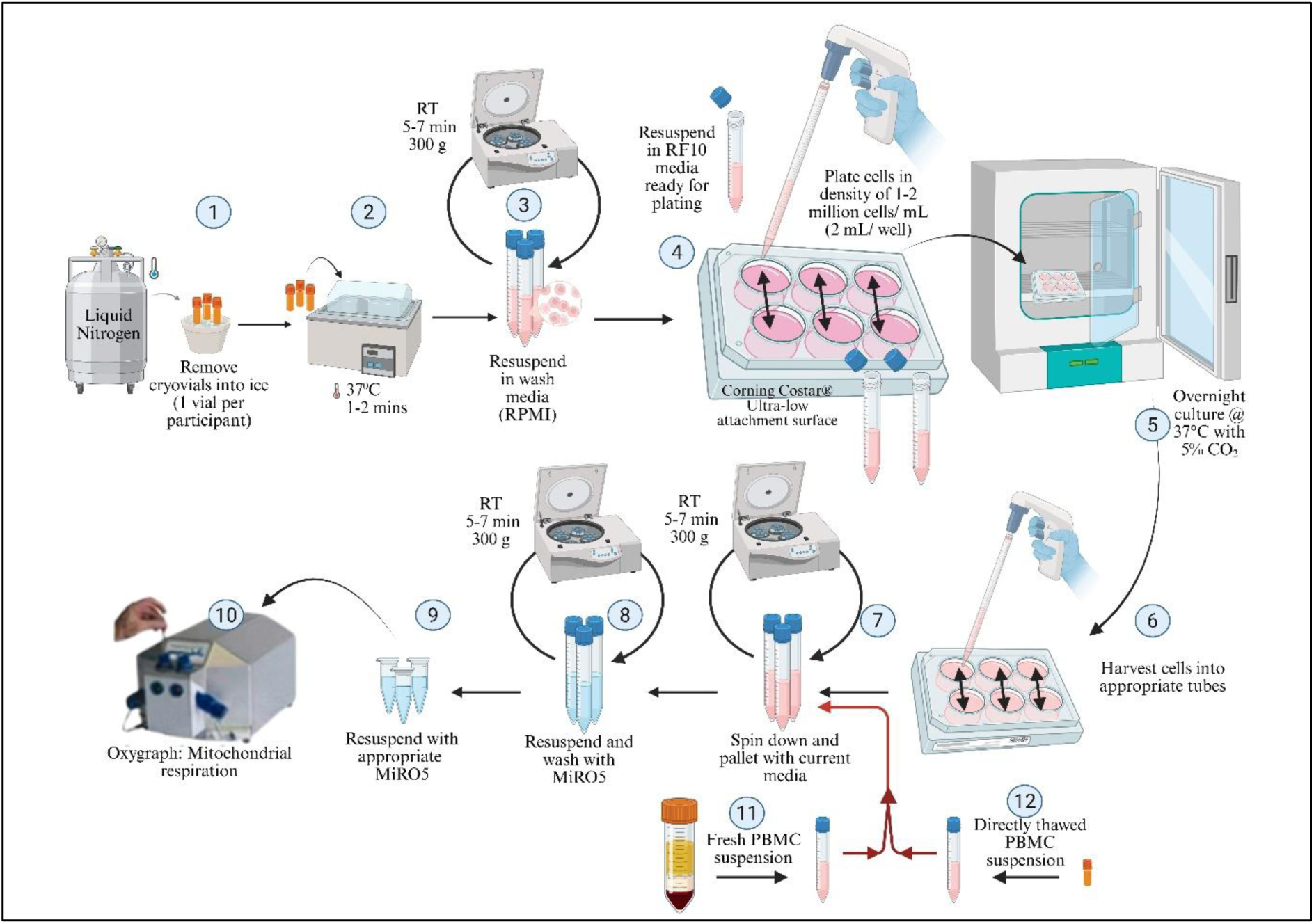
Overview of PBMC preparation workflow for oxygraph analysis. Steps 1–5 represent the thawing, washing, and plating of PBMCs for overnight culture in ultra-low attachment plates. Steps 6–9 describe the harvesting of cultured cells and their subsequent washing and resuspension in MiR05 buffer to prepare for analysis. Step 10 corresponds to mitochondrial respiration measurements using the oxygraph. Steps 11–12 indicate parallel processing of fresh PBMCs and directly thawed PBMCs, which enter the workflow at the preparation stage prior to oxygraph analysis.

**Table 1:** Reagents and media used for the preparation and culture of primary human cells and cell lines in this study.

| Name | Formular | Brand | Use |
|---|---|---|---|
| RPMI (Gibco™ RPMI 1640 medium) | RPMI 1640 +/- Phenol Red | Thermo Fisher Scientific | Wash media |
| RØ | RPMI supplemented with 1x Pen/Strep and 1x GlutaMax™-I | Thermo Fisher Scientific | Base media |
| RF10 | RØ + 10% v/v FBS | Thermo Fisher Scientific | Culture media |
| Freezing Medium | 80% v/v FBS +20% v/v DMSO |  | All cell cryopreservation |
| Fetal Bovine Serum (FBS) | heat inactivated at 56°C | Thermo Fisher Scientific | Included in all cell culture media |
| Penicillin/Streptomycin (Gibco®) |  | Thermo Fisher Scientific | unless stated otherwise |
| GlutaMAX™-I (100x, Gibco®) |  | Thermo Fisher Scientific | Used in base media (RØ) |

RØ: RPMI base medium supplemented with antibiotics (penicillin/streptomycin) and GlutaMax™-I (a stabilized form of L-glutamine). RF10: RØ further enriched with 10% Fetal Bovine Serum (FBS) for supporting cell line growth. Pen/Strep: Antibiotics to prevent bacterial contamination. DMSO: Dimethyl sulfoxide, used as a cryoprotectant in freezing media. FBS: Fetal Bovine Serum, provides nutrients and growth factors.

**Table 2:** Participant characteristics.

| Participant characteristics | Mean ± S.E.M |
|---|---|
| Age (years) | 34.0 ± 4.22 |
| Height (cm) | 180.6 ± 4.33 |
| Weight (kg) | 105.1 ±16.48 |
| Body mass index (m/kg<sup>2</sup>) | 29.3 ± 7.93 |

S.E.M. standard error of the mean.

### Mitochondrial respiration

PBMC suspensions for oxygraph analysis, whether fresh or cryopreserved, were processed using identical steps to ensure consistency. Cells were diluted dropwise with prewarmed wash media (RPMI 1640, Table 1) and gently mixed to obtain a homogeneous suspension, with cryopreserved samples fully thawed prior to processing (**Figure 2**).

For overnight-cultured PBMCs, cells were harvested into 15 mL tubes containing wash media. All samples were centrifuged at 300 × g for 5–7 min at room temperature, and the supernatant was carefully removed without disturbing the pellet. Cells were washed twice and resuspended in MiR05 buffer at an appropriate volume for cell counting and downstream analysis (**Figure 2)**.

Mitochondrial respiration was measured using high-resolution respirometry (Oroboros O2k; Oroboros Instruments, Innsbruck, Austria) with a substrate–uncoupler–inhibitor titration (SUIT) protocol (**Figure 3**) [15-17]. Measurements were performed in duplicate using 6–8 × 10⁶ cells per chamber.

**Figure 3:**
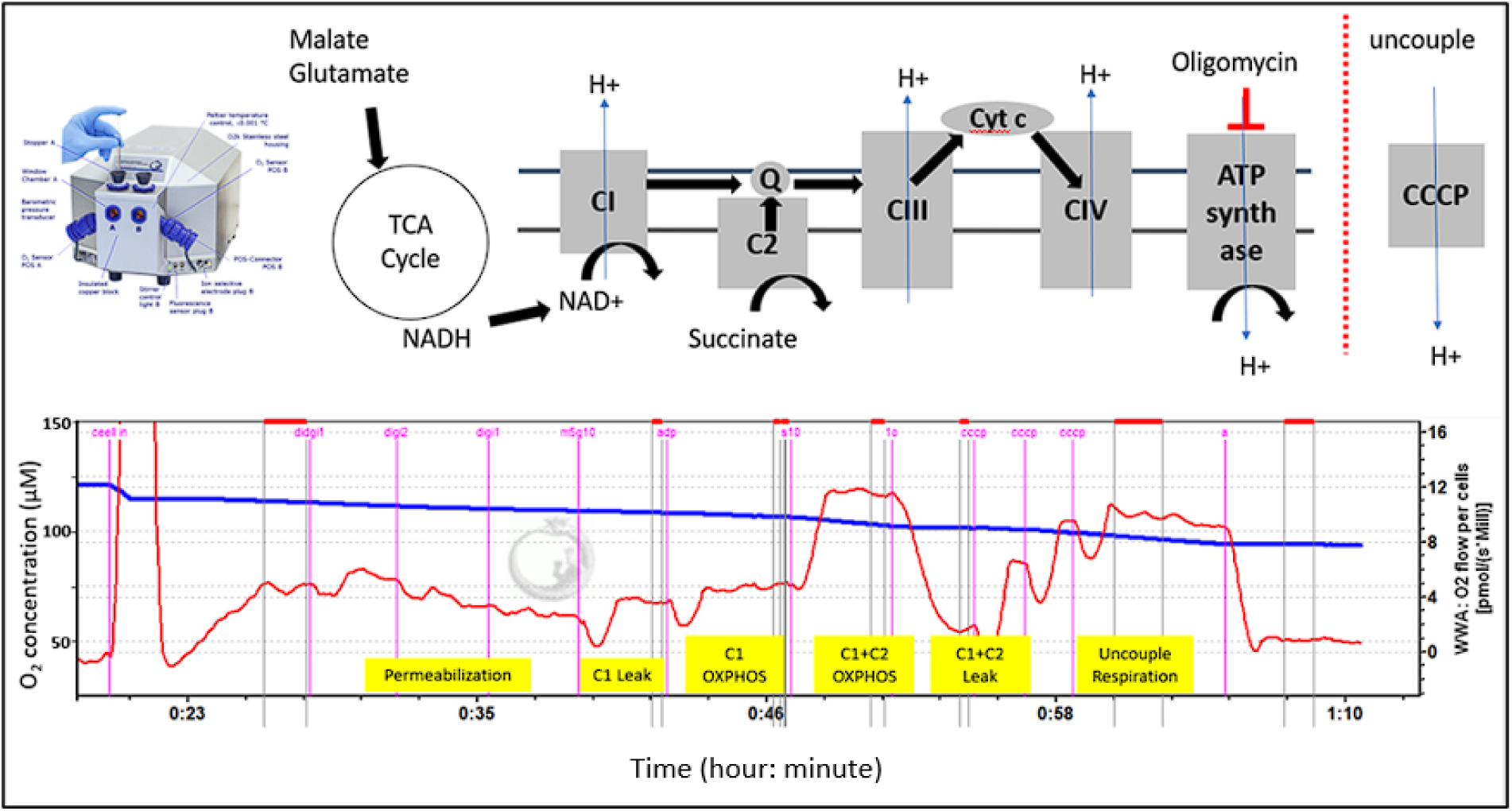
Example demonstration of the mitochondrial respiration assay. Step-by-step schematic illustrating the use of SUIT protocol to measure mitochondrial respiration assay performed using high-resolution respirometry. The workflow includes plasma membrane permeabilisation, sequential substrate–uncoupler–inhibitor titrations, and measurement of defined respiratory states. Representative oxygen flux traces and corresponding quantification of respiration rates generated using DatLab 6.1 software (Oroboros Instruments) are shown.

PBMCs were permeabilised with digitonin (final concentration: 3–4 μM) added to the oxygraph chamber before the respiration titration protocol commenced. Mitochondrial respiration was measured in a range of states representing nonphosphorylating (leak), phosphorylating (oxphos), and uncoupled respiration. Complex I (CI) leak respiration was measured with the addition of malate (2 mM) and pyruvate (5 mM), following which ADP (2.5 mM) was added to measure CI oxphos. Succinate was then added to measure oxphos supported by complex I and II substrates (CI and II oxphos). Oligomycin was then added to inhibit ATP synthase to measure CI and CII leak respiration. Stepwise titrations of 0.5 M CCCP were then added to uncouple respiration from oxidative phosphorylation before respiration was inhibited at complex III (CIII) with antimycin A.

All respiration rates were corrected for non-mitochondrial oxygen consumption using the oxygen consumption rate determined following inhibition of CIII, and the CIV respiration rate was corrected for auto-oxidation in each assay. All mitochondrial respiration rates were recorded and quantified using Datlab 6.1 software (Oroboros Instruments). Data are expressed as respiration rate per million cells for PBMCs.

### Statistical Analysis

Statistical analyses were performed using GraphPad Prism 10 (GraphPad Software, San Diego, CA, USA). Data are presented as mean ± SEM. Comparisons between fresh and frozen PBMC conditions were performed using paired t-tests. Statistical significance was accepted at p < 0.05.

## Results

### Participant characteristics

PBMCs collected from six healthy volunteers were used to determine optimal conditions for downstream assays. Participants had a mean age of 34.0 ± 4.22 years and a mean body mass index (BMI) of 29.28 ± 7.93 kg/m². Each volunteer donated 50–60 mL of whole blood, from which PBMCs were isolated and used for this study.

### PBMC viability between direct thaw and overnight culture

Following revival of PBMCs from cryogenic storage (2 weeks to 1 month), cell viability was compared between two recovery approaches: direct thawing and overnight culture, to determine the most reliable method for downstream assays. Frozen vials were rapidly thawed, transferred into 15 mL of pre-warmed RPMI 1640, and centrifuged at 200 × g for 2 minutes at room temperature to remove DMSO. Cells were washed twice with fresh RPMI and resuspended in 5 mL of complete culture media (RPMI 1640 supplemented with 10% FBS, 1% Penicillin–Streptomycin, and 1% GlutaMAX™). PBMCs were then plated at a density of 2 × 10⁶ cells per well in 24-well plates and either assessed immediately or after an overnight recovery period. Viability analysis revealed no significant difference between directly thawed and overnight-cultured cells (**Table 3**; **Figure 4**), indicating that both approaches provide comparable outcomes and supporting the robustness of the culture workflow for cryopreserved PBMCs.

**Figure 4:**
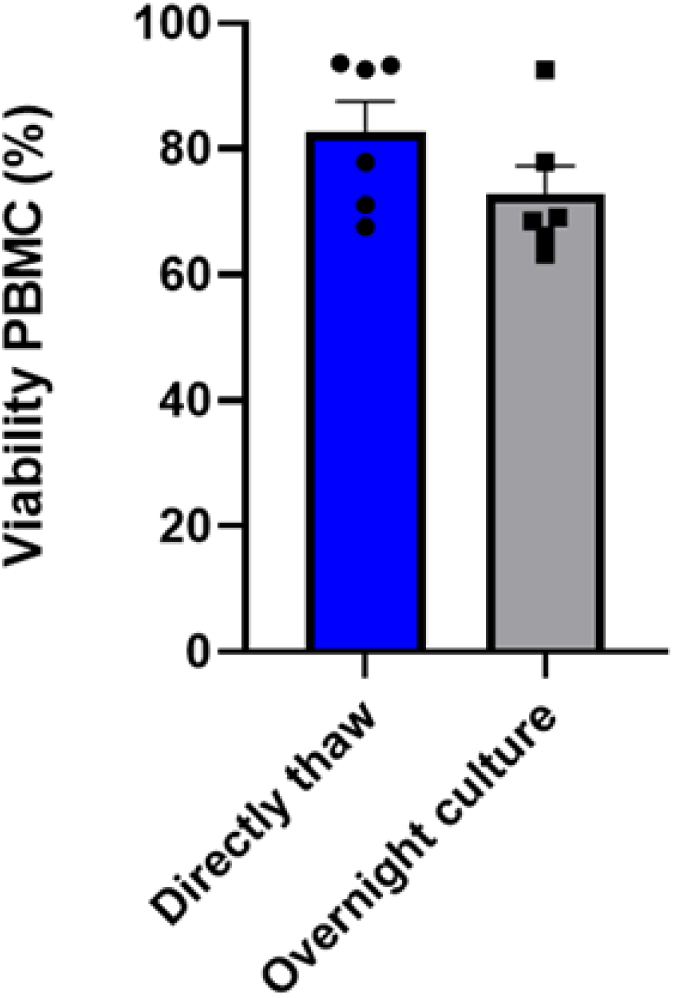
Comparison of PBMC viability between direct thaw and overnight culture conditions. Bars represent mean viability (%) ± standard deviation for six paired samples: Direct thaw (82.7 ± 12.0%) and overnight culture (72.9 ± 10.8%). Individual data points are shown. Although direct thaw exhibited higher mean viability, a paired t-test indicated that this difference was not statistically significant (p = 0.094).

**Table 3:** Participants’ PBMCs viability percentage.

| Participant ID | Directly (%) | Overnight (%) | Delta value |
|---|---|---|---|
| DP1 | 67.6 | 66.2 | 1.4 |
| DP2 | 77.9 | 77.9 | 0 |
| DP3 | 92.6 | 68.5 | 24.1 |
| DP4 | 71.1 | 63.2 | 7.9 |
| DP6 | 93.3 | 92.5 | 0.8 |
| DP7 | 93.6 | 69.1 | 24.5 |
| Mean | 82.7 | 72.9 | 9.8 |
| SD | 12.0 | 10.8 | 1.2 |
| CV% | 14.5 | 14.8 | -0.3 |

SD; Standard deviation. CV%; Percentage of Coefficient of variation.

### Effect of cryopreservation on mitochondrial respiration in human PBMCs

Cryopreservation and thawing can induce membrane and metabolic stress in PBMCs, transiently altering oxidative phosphorylation. Following our comparison of PBMC viability between direct-thaw and overnight-culture conditions (**Figure 4**), we then compared freshly isolated PBMCs with directly thawed and thawed-then-recovered cells to determine whether an overnight recovery step could restore mitochondrial respiration toward fresh profiles.

We hypothesised that culturing cells overnight in fully supplemented RPMI would help minimise these stress effects, normalise nutrient uptake, and provide a more physiologically relevant condition for comparison with freshly isolated PBMCs.

This experimental design enabled us to determine whether cryopreservation affects the mitochondrial phenotype, informing whether PBMCs can be frozen and subsequently used for mitochondrial function measurements. In addition, it allowed us to assess changes in mitochondrial respiration associated with freezing and recovery, independent of cell viability, which remained comparable across all conditions (**Figure 5**).

**Figure 5:**
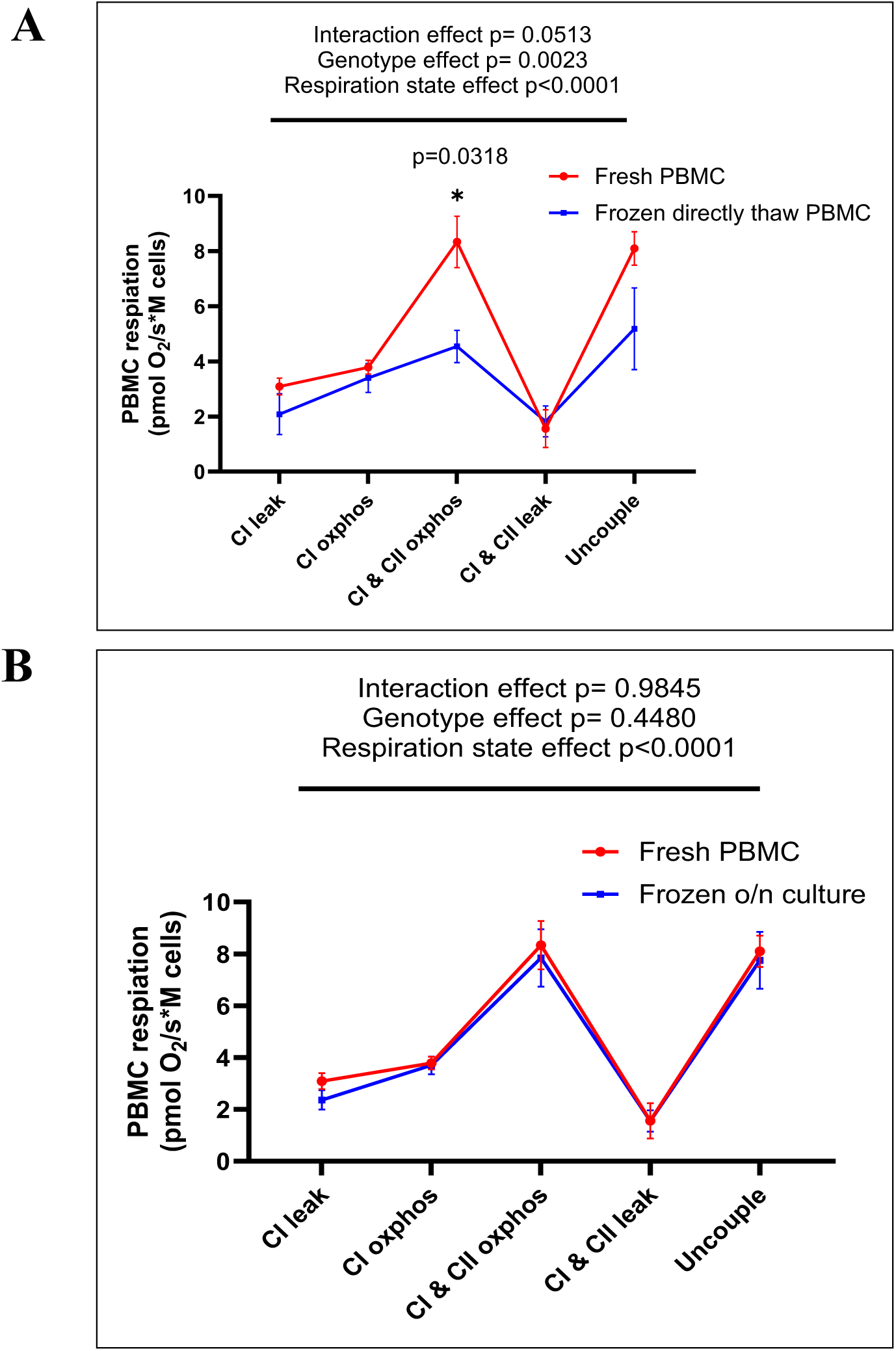
High-resolution respirometry of PBMCs showing cryopreservation-associated changes in oxidative phosphorylation. (A) Comparison of freshly isolated PBMCs and cryopreserved PBMCs analysed immediately after thawing (direct-thaw) demonstrates attenuated basal and ATP-coupled oxygen consumption rates (CI & CII OXPHOS, p = 0.03), along with reduced FCCP-stimulated maximal respiration and spare respiratory capacity (n = 6). (B) Comparison of freshly isolated PBMCs and cryopreserved PBMCs following overnight recovery in supplemented RPMI shows partial restoration of oxidative phosphorylation metrics toward fresh profiles (n = 6). All assays were performed in matched donor pairs across conditions to minimise inter-individual variability.

**Figure 6:**
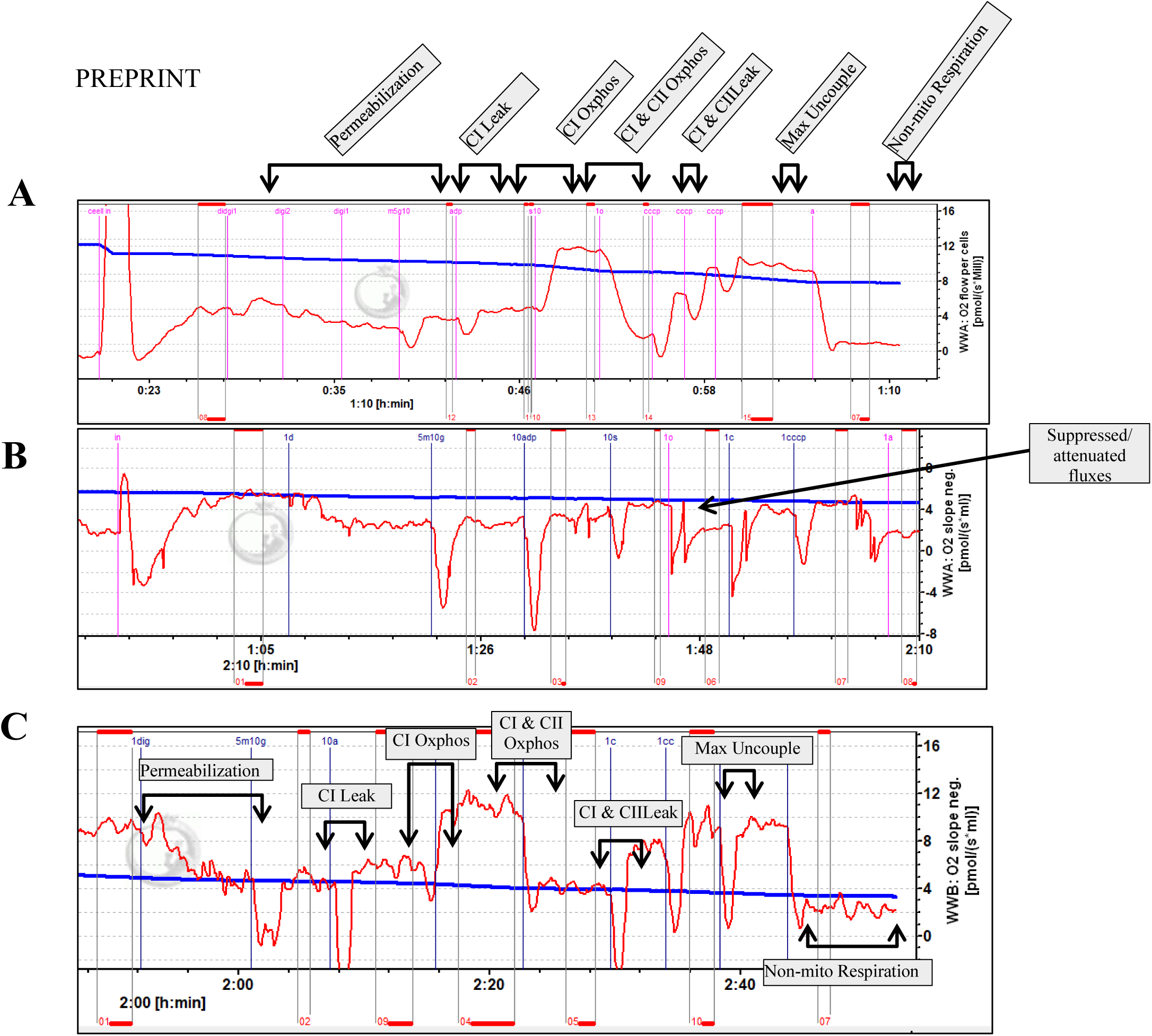
Representative high-resolution respirometry traces of PBMCs under fresh and cryopreserved conditions. (A) Freshly isolated PBMCs assayed immediately after isolation, exhibiting stable oxygen consumption rate (OCR) traces. (B) Cryopreserved PBMCs analysed immediately following thawing (direct-thaw), showing reduced respiratory oxygen fluxes. (C) Cryopreserved PBMCs analysed following overnight recovery in supplemented RPMI, demonstrating restoration of OCR profiles toward fresh conditions. Sequential additions indicate defined respiratory states, including oligomycin (ATP synthase inhibition), FCCP (uncoupling), and rotenone/antimycin A (complex I/III inhibition). Oxygen fluxes were normalised to cell number, and non-mitochondrial respiration was subtracted to derive net mitochondrial respiration. CI = Complex I; CII = Complex II; OXPHOS = oxidative phosphorylation; Maximal uncoupled = maximal uncoupled respiration; Non-mito respiration = non-mitochondrial sources of oxygen consumption.

## Discussion

In this study, we established a workflow incorporating PBMC isolation, cryopreservation, post-thaw recovery, and high-resolution respirometry to evaluate the feasibility of measuring mitochondrial respiration in frozen human PBMCs. Our findings demonstrate that cryopreserved PBMCs can be reliably used for high-resolution respirometry when an overnight recovery period is included following thawing. This is particularly relevant given that cryopreservation is known to induce transient mitochondrial stress, reduce oxidative phosphorylation efficiency, and alter respiratory fluxes across a range of primary cell types[13, 18, 19].

High-resolution respirometry is widely used to assess mitochondrial function in tissues such as skeletal muscle, liver, brain, and heart. However, validated approaches for frozen PBMCs remain limited, with relatively few studies directly addressing methodological constraints in these cells [20]. This limitation has restricted the use of PBMCs in large-scale or multi-centre human studies, where immediate processing of fresh samples is often impractical.

A recognised challenge in PBMC-based respirometry is cellular heterogeneity and platelet contamination, both of which can influence oxygen consumption rate (OCR) measurements. Platelets, which have a high mitochondrial density, can co-purify with PBMCs and potentially lead to overestimation of mitochondrial respiration if not adequately controlled [11]. Despite these challenges, previous studies have reported that cryopreserved PBMCs retain approximately 90–95% of the maximal respiratory capacity observed in freshly isolated cells cells [20]. Consistent with these observations, our data indicate that mitochondrial respiratory capacity is largely preserved following cryopreservation when an appropriate recovery period is applied [21].

Direct analysis of PBMCs immediately after thawing revealed attenuated oxidative phosphorylation and reduced maximal respiratory capacity, consistent with acute cryopreservation-associated mitochondrial suppression. Importantly, these changes were not accompanied by reduced cell viability, suggesting that cryopreservation primarily induces functional, rather than cytotoxic, alterations. Following overnight recovery in fully supplemented culture conditions, mitochondrial respiratory profiles closely approximated those of freshly isolated PBMCs, supporting the inclusion of a recovery step as a critical component of the experimental workflow.

This study has several limitations. The sample size was modest (n = 6), and all participants were healthy young adults, which may limit the generalisability of these findings to older individuals or clinical populations. In addition, PBMC heterogeneity was not further characterised, and differences in cell subset composition may contribute to variability in mitochondrial measurements. Future studies should address these factors and assess the applicability of this workflow across diverse cohorts and disease states.

Overall, these findings validate a practical and reproducible framework for assessing mitochondrial respiration in cryopreserved PBMCs using high-resolution respirometry. Incorporation of an overnight recovery period overcomes a key limitation associated with reliance on freshly isolated cells, enabling sample batching, multi-site study designs, and retrospective analysis of biobanked material. This approach broadens the applicability of PBMC-based mitochondrial phenotyping in human research.

## Data Availability

The datasets generated and analysed during the current study are available from Robert Douglas ’Atiola upon reasonable request.

## References

1. Kühlbrandt, W., Structure and function of mitochondrial membrane protein complexes. BMC Biology, 2015. 13(1): p. 89.

2. Mitchell, P., Chemiosmotic coupling in oxidative and photosynthetic phosphorylation. 1Scc. Biochim Biophys Acta, 2011. 1807(12): p. 1507–38.

3. Gosling, A.L., et al., Mitochondrial genetic variation and gout in Maori and Pacific people living in Aotearoa New Zealand. Ann Rheum Dis, 2018. 77(4): p. 571–578.

4. Lee, D.W., et al., Loss of a conserved tyrosine residue of cytochrome b induces reactive oxygen species production by cytochrome bc1. J Biol Chem, 2011. 286(20): p. 18139–48.

5. Rosa, H.S., et al., A case for measuring both cellular and cell-free mitochondrial DNA as a disease biomarker in human blood. The FASEB Journal, 2020. 34(9): p. 12278–12288.

6. Turrens, J.F., Mitochondrial formation of reactive oxygen species. J Physiol, 2003. 552(Pt 2): p. 335–44.

7. Anderson, E.J., et al., Substrate-specific derangements in mitochondrial metabolism and redox balance in the atrium of the type 2 diabetic human heart. J Am Coll Cardiol, 2009. 54(20): p. 1891–8.

8. Anderson, E.J., et al., Increased propensity for cell death in diabetic human heart is mediated by mitochondrial-dependent pathways. Am J Physiol Heart Circ Physiol, 2011. 300(1): p. H118–24.

9. Mohlke, K.L., et al., Mitochondrial polymorphisms and susceptibility to type 2 diabetes-related traits in Finns. 2005. 118: p. 245–254.

10. Shen, F.C., et al., Mitochondrial haplogroups have a better correlation to insulin requirement than nuclear genetic variants for type 2 diabetes mellitus in Taiwanese individuals. 2022. 13(1): p. 201–208.

11. Sharma, E., et al., Overview of methods that determine mitochondrial function in human disease. Metabolism, 2025. 170: p. 156300.

12. Van Bergen, N.J., et al., Oxidative phosphorylation measurement in cell lines and tissues. Mitochondrion, 2014. 15: p. 24–33.

13. Osto, C., et al., Measuring mitochondrial respiration in previously frozen biological samples. Current Protocols in Cell Biology, 2020. 89(1): p. e116.

14. Acin-Perez, R., et al., A novel approach to measure mitochondrial respiration in frozen biological samples. EMBO Journal, 2020. 39(13).

15. Timón-Gómez, A., et al., Analysis of mitochondrial respiratory pathway and coupling control by substrateuncoupler-inhibitor titration reference protocols. 2024, Zenodo.

16. Kuznetsov, A.V., et al., Analysis of mitochondrial function in situ in permeabilized muscle fibers, tissues and cells. Nat Protoc, 2008. 3(6): p. 965–76.

17. Hedges, C.P., et al., Peripheral blood mononuclear cells do not reffect skeletal muscle mitochondrial function or adaptation to high-intensity interval training in healthy young men. 2019. 126(2): p. 454–461.

18. Acin-Perez, R., et al., A novel approach to measure mitochondrial respiration in frozen biological samples. The EMBO Journal, 2020. 39(13): p. e104073.

19. Hope, C.M., et al., Optimization of Blood Handling and Peripheral Blood Mononuclear Cell Cryopreservation of Low Cell Number Samples. Int J Mol Sci, 2021. 22(17).

20. Sharma, E., et al., Overview of methods that determine mitochondrial function in human disease. Metabolism, 2025: p. 156300.

21. *Product information and technical documentation*, in *STEMCELL Technologies Inc*., STEMCELL Technologies.

